# Effect of testosterone therapy on breast tissue composition and mammographic breast density in trans masculine individuals

**DOI:** 10.1101/2024.01.09.24300987

**Authors:** Yujing J Heng, Gabrielle M Baker, Valerie J Fein-Zachary, Yaileen D Guzman-Arocho, Vanessa C Bret-Mounet, Erica S Massicott, Sy Gitin, Paul Russo, Adam M Tobias, Richard A Bartlett, Gopal Varma, Despina Kontos, Lusine Yaghjyan, Michael S Irwig, Jennifer E Potter, Gerburg M Wulf

**Affiliations:** Department of Pathology, Beth Israel Deaconess Medical Center, Harvard Medical School, Boston, MA; Department of Radiology, Beth Israel Deaconess Medical Center, Harvard Medical School, Boston, MA; The Fenway Institute, Boston, MA; Department of Surgery, Beth Israel Deaconess Medical Center, Harvard Medical School, Boston, MA, USA; Department of Medicine, Beth Israel Deaconess Medical Center, Harvard Medical School, Boston, MA; Perelman School of Medicine, University of Pennsylvania, Philadelphia, PA; University of Florida, College of Public Health and Health Professions and College of Medicine, Department of Epidemiology, Gainesville, FL

**Keywords:** gender-affirming hormones, transgender health disparities, breast cancer risk, trans men, non binary people, hormone replacement therapy, top surgery

## Abstract

**Objective:** Determine the association between TT and breast tissue composition and breast tissue density in trans masculine individuals (TMIs).

**Design:** This is a cross-sectional study.

**Setting:** TMIs (n=444) underwent chest-contouring surgeries to treat their gender dysphoria between 2013 and 2019 at an urban medical center.

**Participants:** Of the 444 TMIs, 425 had pathology images analyzed by our deep-learning algorithm to extract breast tissue composition. A subset of 42/444 TMIs had mammography prior to surgery; mammography files were available for 25/42 TMIs and analyzed using a breast density software, LIBRA.

**Main Outcome(s) and Measure(s):** The first outcome was the association of duration of TT and breast tissue composition assessed by pathologists (categories of lobular atrophy and stromal composition) or by our algorithm (% epithelium, % fibrous stroma, and % fat). The second outcome is the association of TT and breast density as assessed by a radiologist (categorical variable) or by LIBRA (percent density, absolute dense area, and absolute non-dense area).

**Results:** Length of TT was associated with increasing degrees of lobular atrophy (*p*<0.001) but not fibrous content (*p*=0.821) when assessed by the pathologists. Every six months of TT was associated with decreased amounts of both epithelium (exp(β)=0.97, 95% CI 0.95-0.98, adj *p*=0.005) and stroma (exp(β)=0.99, 95% CI 0.98-1.00, adj *p*=0.051), but not fat (exp(β)=1.01, 95%CI 0.98-1.05, *p*=0.394) in fully adjusted models. There was no association between TT and radiologist’s breast density assessment (*p*=0.575) or LIBRA measurements (*p*>0.05).

**Conclusions:** TT decreases breast epithelium and fibrous stroma, thus potentially reducing the breast cancer risk of TMIs. Further studies are warranted to elucidate the effect of TT on breast density and breast cancer risk.

**Summary Box:**

- Very little is known about the effect of gender-affirming testosterone therapy on cancer risks, such as breast cancer.
- Epidemiological studies had different conclusions about the association between testosterone and breast cancer in cisgender women (positive association) and trans masculine individuals (inverse association).
- More laboratory-based research are needed to understand the effect of testosterone on breast cancer risk in the understudied trans masculine population.
- Our study provides quantitative histological evidence to support prior epidemiological reports that testosterone may reduce breast cancer risk in trans masculine individuals.

## Introduction

About 65% of trans masculine individuals (TMIs) pursue testosterone therapy (TT) to treat their gender dysphoria (1). TMIs are defined in this paper as individuals who were born female and identify as trans men or non-binary people. The breast is highly sensitive to sex hormones. Normal breast proliferation is regulated by the balance between the stimulating effects of estrogens and the inhibitory effects of testosterone. TT increases the baseline testosterone levels of TMIs by ≥10-fold to achieve levels comparable to cisgender men (2). The extent to which TT affects breast tissue and breast cancer (BC) risk remains unclear.

While TT has been explored to prevent and/or treat BC in cisgender women (3–6), epidemiological studies in cisgender women reported an association between high circulating testosterone levels and increased BC risk (7–12). One explanation for that phenomenon is that testosterone can be aromatized to estradiol, contributing to breast cell proliferation and tumorigenesis (13). In contrast, three epidemiological studies that examined BC incidence in trans people concluded that trans men do not have an increased BC risk compared to cisgender females (14–16).

Most trans masculine breast-related studies to date are histopathological reviews of breast tissues obtained after chest-contouring surgeries. We and others have shown that TT has a profound effect on non-cancer breast morphology (17–23). Our group observed that alterations in breast histology, in particular, higher degrees of lobular atrophy, were evident after ≥12 months of TT (23). We also subsequently reported that that Toker cell hyperplasia, a histological feature with unclear clinical significance, was more frequently observed in TMIs compared to cisgender women (24).

Mammographic breast density reflects the relative amounts of breast fibroglandular (epithelium and stroma) and fat. High mammographic breast density is a strong BC risk factor for cisgender women (25). We previously developed a computational pathology algorithm to assess non-cancer breast tissue composition (26–29). Using our algorithm, women with more breast epithelium (highest quartile) had higher subsequent BC risk compared with women in the lowest quartile (28). Hormone-related BC risk factors such as reproductive and early body weight were also linked to changes in breast tissue composition (26,27). Thus, it is important to understand the effect of TT on breast tissue composition in order to understand trans masculine BC risk.

To address these knowledge gaps, we investigated the association of TT and breast tissue composition in 425 TMIs. In a subset of 42 subjects who had mammograms, we explored the relationship between TT and mammographic breast density, and performed radiology-pathology correlations.

## Materials and Methods

### Study Subjects

This cross-sectional study initially included 444 TMIs who had chest-contouring surgery at an urban medical center between 2013 and 2019 (23,30). Clinical data were retrieved from medical record, including age at surgery, race/ethnicity, family history of BC, parity, oophorectomy status at time of surgery, body mass index (BMI), alcohol consumption, TT regimen, duration of TT (months), and whether they bound their chest. We estimated the duration of TT at time of surgery in one of two ways depending on data availability: 1) by calculating the number of months between date of the first TT prescription and date of surgery, or 2) by combining the subject’s verbal recount of how long they had been receiving TT at their pre-surgical consult and the time between that pre-surgical consult and date of surgery. This study was IRB approved. Figure 1 summarizes the histological, radiological, and quantitative data used in this study. The study was approved by the BIDMC Institutional Review Board (2018P000814).

**Figure 1.**
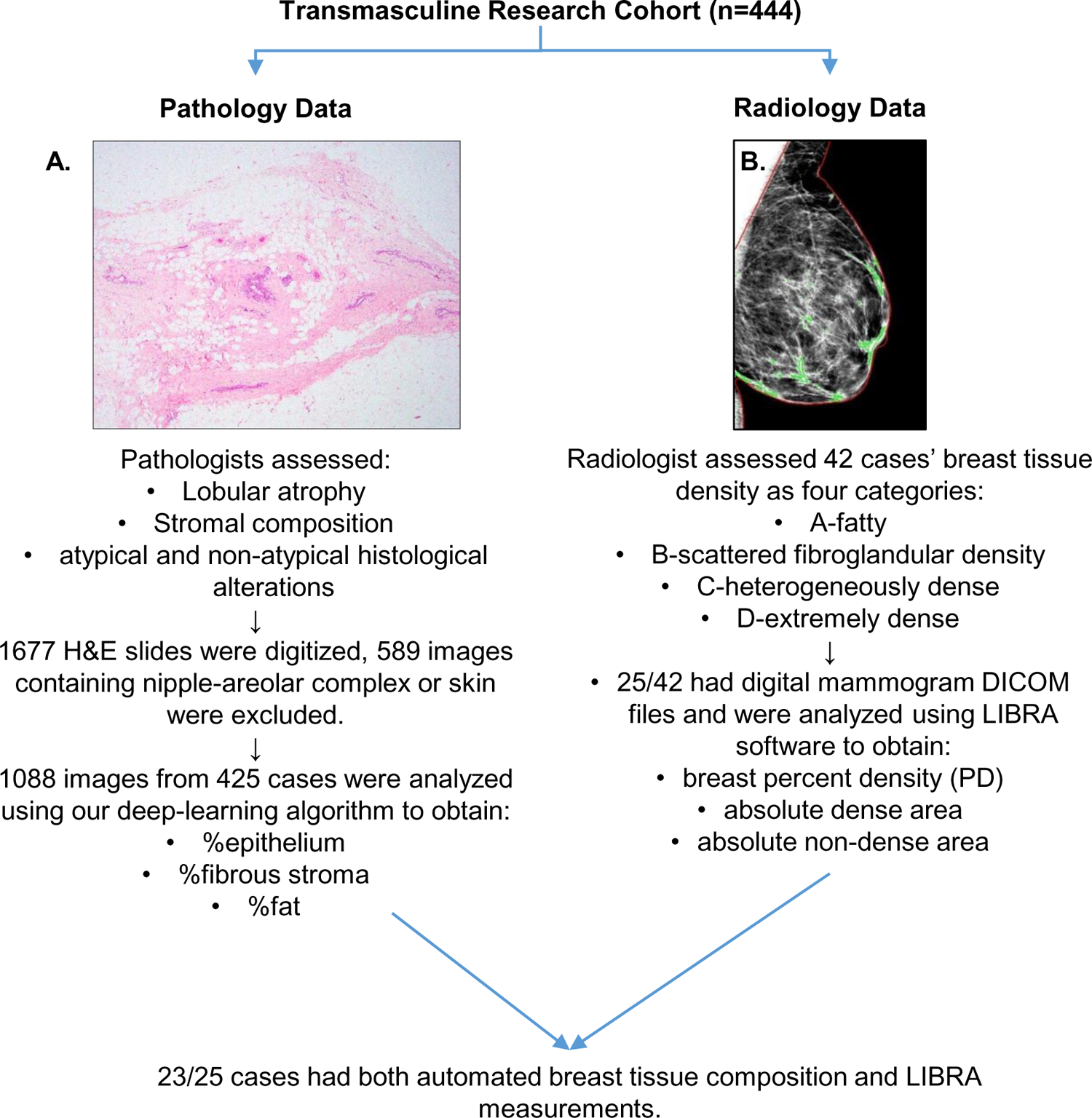
Summary of the histological, radiological, and quantitative data used for each part of this study. (**A**) Breast histology image from a subject who had been using testosterone therapy for 4.2 years at the time of chest-contouring surgery. Pathologists assessed the breast tissue as demonstrating a moderate degree of lobular atrophy with mixed fatty and fibrous stroma. Our algorithm quantified this breast tissue as containing 2.8% epithelium, 53.1% fibrous stroma, and 44.1% fat. (**B**) Corresponding mammogram from the same subject taken two months prior to surgery. The radiologist classified this case as B-scattered fibroglandular density. The Laboratory for Individualized Breast Radiodensity Assessment (LIBRA) software estimated the breast percent density as 1.9%. Digital imaging and communications in medicine, DICOM.

### Pathological review and automated breast tissue composition

The pathology department’s grossing protocol for chest-contouring specimens was to sample each quadrant of the breast parenchyma, and submit two blocks per breast. Additional sections were submitted if the nipple or skin was present, and if a gross lesion or atypia was identified (23). For each of the 444 cases, H&E-stained slides were reviewed by two pathologists (GMB and YDG). Each case was assessed for 1) degree of lobular atrophy classified as minimal, mild, moderate, or marked (Figure 2A), 2) stromal composition classified as predominantly fatty, mixed fatty and fibrous, and predominantly fibrous (Figure 2C), and 3) atypical and non-atypical histological alterations (23). Atypical breast lesions included ductal carcinoma *in situ* (DCIS), lobular carcinoma *in situ*, atypical ductal hyperplasia (ADH), atypical lobular hyperplasia (ALH), and flat epithelial atypia. Slides (*n*=1677) were digitized at 20× using the Pannoramic Scan P150 (3DHISTECH Ltd, Budapest, Hungary).

**Figure 2.**
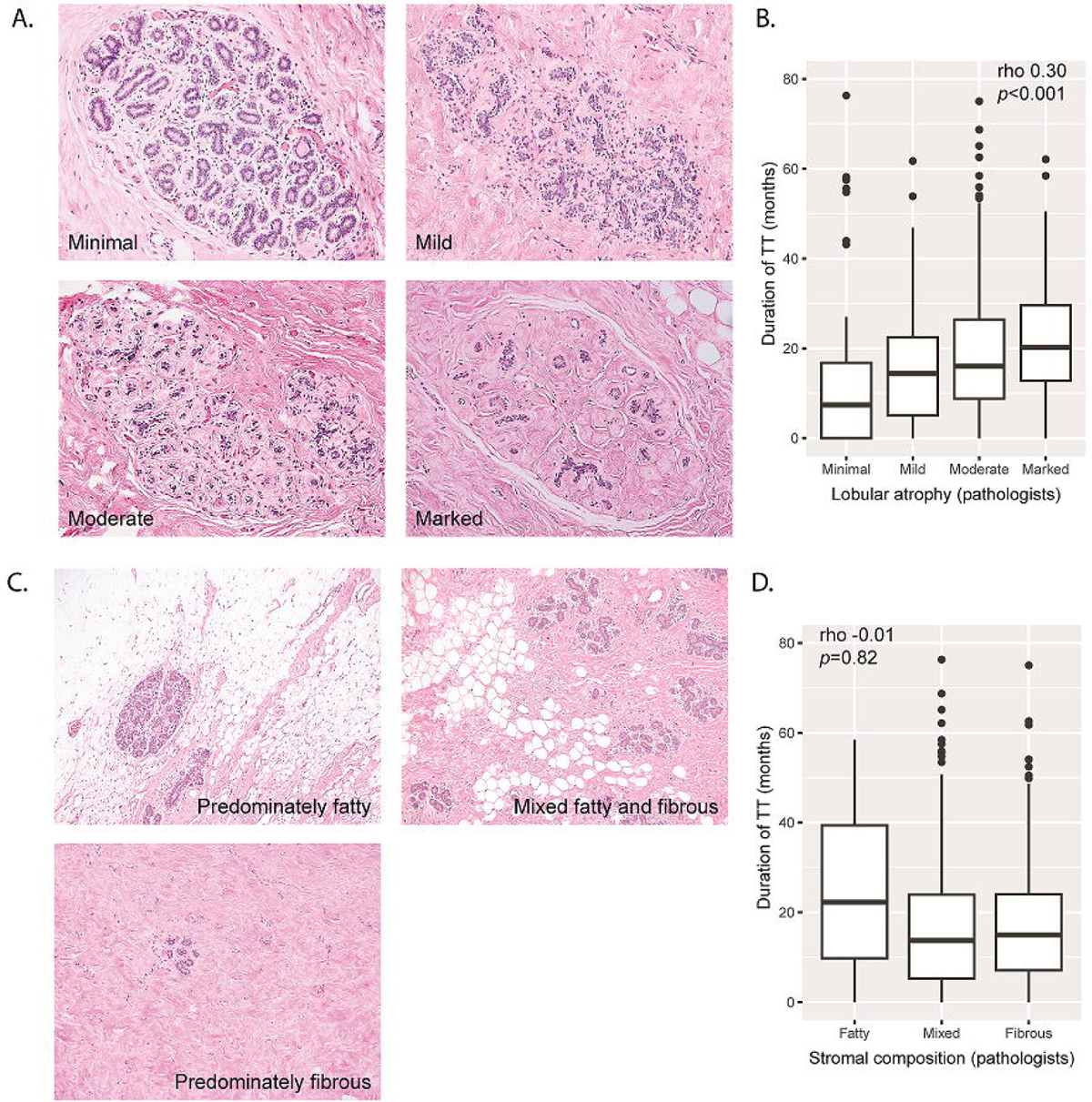
(**A**) Lobular atrophy was assessed by the pathologists using four categories. (**B**) The duration of testosterone therapy (TT) significantly correlated with increasing degrees of lobular atrophy. (**C**) Stromal composition was assessed by the pathologists using three categories. (**D**) There was no correlation between duration of TT and stromal composition as assessed by the pathologists.

We previously developed a deep-learning algorithm to segment breast histological images into epithelium, fibrous stroma, and fat (28). As this algorithm was not developed using images containing nipple-areolar complex or skin, it was likely to erroneously classify pixels containing nipple-areolar complex or skin as breast epithelium. Hence, we excluded images with nipple-areolar complex or skin (*n*=589) and applied the algorithm to the remaining 1,088 images from 425 transmasculine subjects. The number of pixels classified as epithelium, fibrous stroma, or fat, were summed across the images of each subject, divided by the total number of tissue pixels detected across all images, and expressed as a percentage (%). Automated breast tissue composition hereafter refers to quantitative % epithelium, % fibrous stroma, and % fat obtained using histological images and our algorithm.

### Radiological assessment and quantitative measures of mammographic breast density

A subset of 42 out of 444 (9.5%) TMIs had mammograms prior to surgery. Additional clinical data were retrieved to reflect oophorectomy status, duration of TT, and chest binding at the time of mammogram. BMI at time of mammogram was not available. The radiologist (VJF) assessed breast tissue density by classifying the densest area as A-fatty, B-scattered fibroglandular density, C-heterogeneously dense, or D-extremely dense.

Digital mammography files in digital imaging and communications in medicine (DICOM) format were available for 25 out of 42 subjects (59.5%). DICOM files for the other 17 subjects were unavailable as their mammograms were conducted elsewhere. DICOM files were processed using the fully automated, publicly available Laboratory for Individualized Breast Radiodensity Assessment (LIBRA) software (31). LIBRA measurements correlate to Cumulus, an established research software, and Volpara, a commercially available software (32). LIBRA identifies the breast region on the mammogram, partitions the breast into gray-level intensity clusters which are then aggregated into the final dense tissue segmentation, and calculates the area of dense pixels to estimate the total absolute dense area. Breast percent density (PD) is obtained by normalizing the absolute dense area by the total breast area (31–33). Absolute non-dense area is calculated by subtracting absolute dense area from total breast area. Since subjects can have up to two mediolateral oblique views per breast, each LIBRA measurement was averaged across multiples DICOM files of the right and left breast. LIBRA measurements hereafter refer to the quantitative values of PD, absolute dense area, and absolute non-dense area.

### Statistical analysis

Mann-Whitney or Fisher’s exact test was used to compare clinical characteristics. Spearman’s rho was used to describe the correlations between pathologists’ assessments and automated breast tissue composition measures, overall and stratified by TT. Spearman’s rho correlated the duration of TT (months; continuous variable) and the pathologists’ assessments. Linear regression modeled the relationship between every six months of TT and automated breast tissue composition (natural log-transformed), adjusting for: (1) age and year of surgery (model 1); or (2) age, year of surgery, race/ethnicity, BMI at surgery, chest binding, and oophorectomy (model 2). To control for differences in TT dosages, we additionally controlled for estimated weekly testosterone (mg) to clarify the effect of dose on tissue composition (model 3). Optimal TT dosages are not solely determined by circulating plasma levels but are frequently adjusted for patients to achieve their desired masculinizing effect. TMIs may receive daily, weekly, or biweekly TT, depending on the mode of administration (transdermal gel/patch, subcutaneous pellet implant, subcutaneous or intra-muscular injection) (30). We also performed analyses stratified by BMI (normal weight BMI<25, overweight BMI ≥25 and <30, and obese BMI≥30), and among nulliparous subjects. To improve the interpretability of the beta coefficient, we back transformed the natural log beta coefficients.

For 42 subjects who had mammography prior to their surgeries, we used spearman’s rho to explore the correlation between TT and breast tissue density assessed by the radiologist, and stratified by age (<40 and ≥40 years old). Among the 25 subjects with DICOM files, we correlated the radiologist’s assessment and LIBRA measurements using Spearman’s rho. The association of TT duration (per six months) with each LIBRA measurement (natural log-transformed) was assessed using linear regression (without adjustment and adjusted for age and BMI at surgery); we also stratified by BMI (≤25 and >25) and restricting to nulliparous subjects.

Twenty three out of 25 subjects had both histological images and DICOM files allowing for correlative analyses using Spearman’s rho. Sensitivity analysis was conducted by restricting to 17 out of 23 individuals who had surgeries within six months after their mammography. Analyses were conducted using R. The level of significance for all statistical tests (2-sided) was *p*<0.05.

## Results

### Subjects with pathology data

The age range of 425 TMIs who had breast tissue composition data was 18 to 61 years old (median=25), and 326 TMIs (76.7%) were White. Three hundred and fifty-seven subjects used TT (84.0%) and 38.9% used TT for one to two years (Table 1). TT users were two years younger (*p*=0.003), and more likely to bind their chest (*p*=0.03) compared to non-users. Atypical breast lesions were only detected among TT users (*p*=0.03): 1 case with DCIS, 5 cases with ADH, 2 cases with ALD, and 1 case had both ADH and ALH. Those who did not use TT were more likely to consume alcohol (*p*=0.01). There was no difference in family history of BC, parity, oophorectomy status, or BMI, between TT users and non-users (*p*>0.05; Table 1).

**Table 1.** Characteristics of the 425 transmasculine subjects pathology data, stratified by testosterone therapy (TT) use.

|  | Used TT<br>*n*=357 | Did not use TT<br>*n*=68 | *p* value |
| --- | --- | --- | --- |
| **Age at surgery, median [IQR]** | 25.0 [21.0, 29.0] | 27.0 [23.8, 32.2] | 0.003<sup>a</sup> |
| **Race/ethnicity, *n* (%)** |  |  | 0.47<sup>b</sup> |
| White | 277 (77.6) | 49 (72.1) |  |
| Black or African American | 30 (8.4) | 6 (8.8) |  |
| Asian | 20 (5.6) | 1 (1.5) |  |
| Multiracial | 18 (5.0) | 4 (5.9) |  |
| Native American/Pacific Islander | 2 (0.6) | 1 (1.5) |  |
| Unspecified | 10 (2.8) | 7 (10.3) |  |
| **Family history of breast cancer, *n* (%)** |  |  | 0.63<sup>b</sup> |
| Yes | 94 (26.3) | 18 (26.5) |  |
| No | 207 (58.0) | 34 (50.0) |  |
| Not reported | 56 (15.7) | 16 (23.5) |  |
| **Parity, *n* (%)** |  |  | 0.41<sup>b</sup> |
| Parous | 8 (2.2) | 3 (4.4) |  |
| Nulliparous | 142 (39.8) | 29 (42.6) |  |
| Not reported | 207 (58.0) | 36 (52.9) |  |
| **Oophorectomy prior to surgery, *n* (%)** |  |  | 0.22<sup>b</sup> |
| Yes | 20 (5.6) | 1 (1.5) |  |
| No | 337 (94.4) | 67 (98.5) |  |
| **Breast lesions, *n* (%)** |  |  | 0.03<sup>b</sup> |
| Atypical lesions | 9 (2.5) | 0 (0.0) |  |
| Benign lesions | 290 (81.2) | 64 (94.1) |  |
| None | 58 (16.2) | 4 (5.9) |  |
| **BMI at surgery, median [IQR]** | 25.8 [23.2, 29.9] | 25.4 [23.0, 29.5] | 0.57<sup>a</sup> |
| **Alcohol consumption, *n* (%)** |  |  | 0.01<sup>b</sup> |
| Current | 186 (52.1) | 48 (70.6) |  |
| Never | 159 (44.5) | 20 (29.4) |  |
| Not reported | 12 (3.4) | 0 (0.0) |  |
| **Duration of TT at surgery, *n* (%)** |  |  | - |
| Never | 0 (0.0) | 68 (100.0) |  |
| <1 year | 93 (26.1) | 0 (0.0) |  |
| >=1 to <2 years | 139 (38.9) | 0 (0.0) |  |
| >=2 to <5 years | 97 (27.2) | 0 (0.0) |  |
| >=5 years | 14 (3.9) | 0 (0.0) |  |
| Unspecified duration | 14 (3.9) | 0 (0.0) |  |
| **Chest binding, *n* (%)** |  |  | 0.03<sup>b</sup> |
| Yes | 169 (47.3) | 20 (29.4) |  |
| No | 15 (4.2) | 6 (8.8) |  |
| Not reported | 173 (48.5) | 42 (61.8) |  |

*p*-values were obtained using the <sup>a</sup>Mann-Whitney or <sup>b</sup>Fisher's exact test. Data in "not reported or unspecified" categories were excluded from statistical analysis. Body mass index, BMI; Inter-quartile range, IQR. Percentages may not add up to 100% due to rounding.

### Association of TT and breast tissue composition

The duration of TT use was significantly correlated with increasing degrees of lobular atrophy (*p*<0.001; Figure 2B) but was not correlated with increasing fibrous content in the stroma (*p*=0.82; Figure 2D) as assessed by the pathologists. Algorithm-derived breast tissue composition data were significantly correlated with pathologists’ assessments (all *p*<0.001; Supplementary 1). For every six months of TT use, the amount of breast epithelium decreased by 3% in the fully adjusted model 3 (exp(β)=0.97, 95% CI 0.95-0.98, *p*=0.005; Table 2). The amount of fibrous stroma also decreased by 1% per six months of TT (model 3, exp(β)=0.99, 95% CI 0.98-1.00, *p*=0.05; Table 2). Although % fat increased by 2% for every six months of TT (model 1 exp(β)=1.02, 95% CI 1.00-1.04, *p*=0.01; Table 2), this association was attenuated in models 2 and 3.

**Table 2.** The association of testosterone therapy (per six months duration) and the percentages (%) of each breast tissue region.

| | | *n* | Exp($\beta$) | 95% CI | *p* value |
| --- | --- | --- | --- | --- | --- |
| **% epithelium** |  |  |  |  |  |
|  | Model 1 | 411 | 0.97 | 0.95-0.98 | <0.001 |
|  | Model 2 | 207 | 0.96 | 0.94-0.98 | 0.001 |
|  | Model 3 | 193 | 0.97 | 0.95-0.99 | 0.005 |
| **% fibrous stroma** |  |  |  |  |  |
|  | Model 1 | 411 | 0.99 | 0.98-0.99 | <0.001 |
|  | Model 2 | 207 | 0.99 | 0.98-1.00 | 0.02 |
|  | Model 3 | 193 | 0.99 | 0.98-1.00 | 0.05 |
| **% fat** |  |  |  |  |  |
|  | Model 1 | 411 | 1.02 | 1.00-1.04 | 0.01 |
|  | Model 2 | 207 | 1.02 | 0.99-1.06 | 0.15 |
|  | Model 3 | 193 | 1.01 | 0.98-1.05 | 0.39 |

Model 1 adjusted for age and year of surgery. Model 2 adjusted for age and year of surgery, race/ethnicity, BMI, chest binding, and oophorectomy status. Model 3 adjusted for age and year of surgery, race/ethnicity, BMI, chest binding, oophorectomy status, and estimated weekly testosterone dose. Confidence interval, CI.

There was no association between TT and % fat when stratified by BMI (Supplementary 2). When restricted to nulliparous subjects, % epithelium decreased by 4% per six months of TT (model 3 exp(β)=0.96, 95% CI 0.92-0.99, *p*=0.02; Supplementary 3). Finally, there was no association between % fibrous stroma or % fat with TT among nulliparous subjects (Supplementary 3).

### Subjects with radiology data

The demographics of the 42 subjects who had pre-operative mammography resembled the larger cohort of 425 subjects (Supplementary 4). The median age in this subset was 43 years (range 20 to 61). Six subjects (14.3%) were assessed by the radiologist as having A-fatty breasts, 13 (31.0%) had B-scattered fibroglandular densities, 18 (42.9%) had C-heterogeneously dense breasts, and five (11.9%) had D-extremely dense breasts. The median time between mammogram to surgery was 4.4 months (range 0.4 to 34.9). Subjects with and without available digital mammogram DICOM files were similar with respect to their demographics (Supplementary 4).

### Association of TT and mammographic breast density

There was no correlation between duration of TT at the time of mammography and the radiologist’s breast tissue density assessment (*p*=0.58; Figure 3), and when stratified by age (<40 years old *n*=20, rho=-0.26, *p*=0.27; ≥40 years old *n*=22, rho=0.19, *p*=0.40). Among the 25 subjects with DICOM files, LIBRA measurements significantly correlated with the radiologist’s assessment (all *p*≤0.003; Supplementary 5), validating the LIBRA measurements. There was no association between the duration of TT and any of the LIBRA measurements (*p*>0.05; Supplementary 6), even when stratified by BMI (Supplementary 7) or restricted to nulliparous women (Supplementary 8).

**Figure 3.**
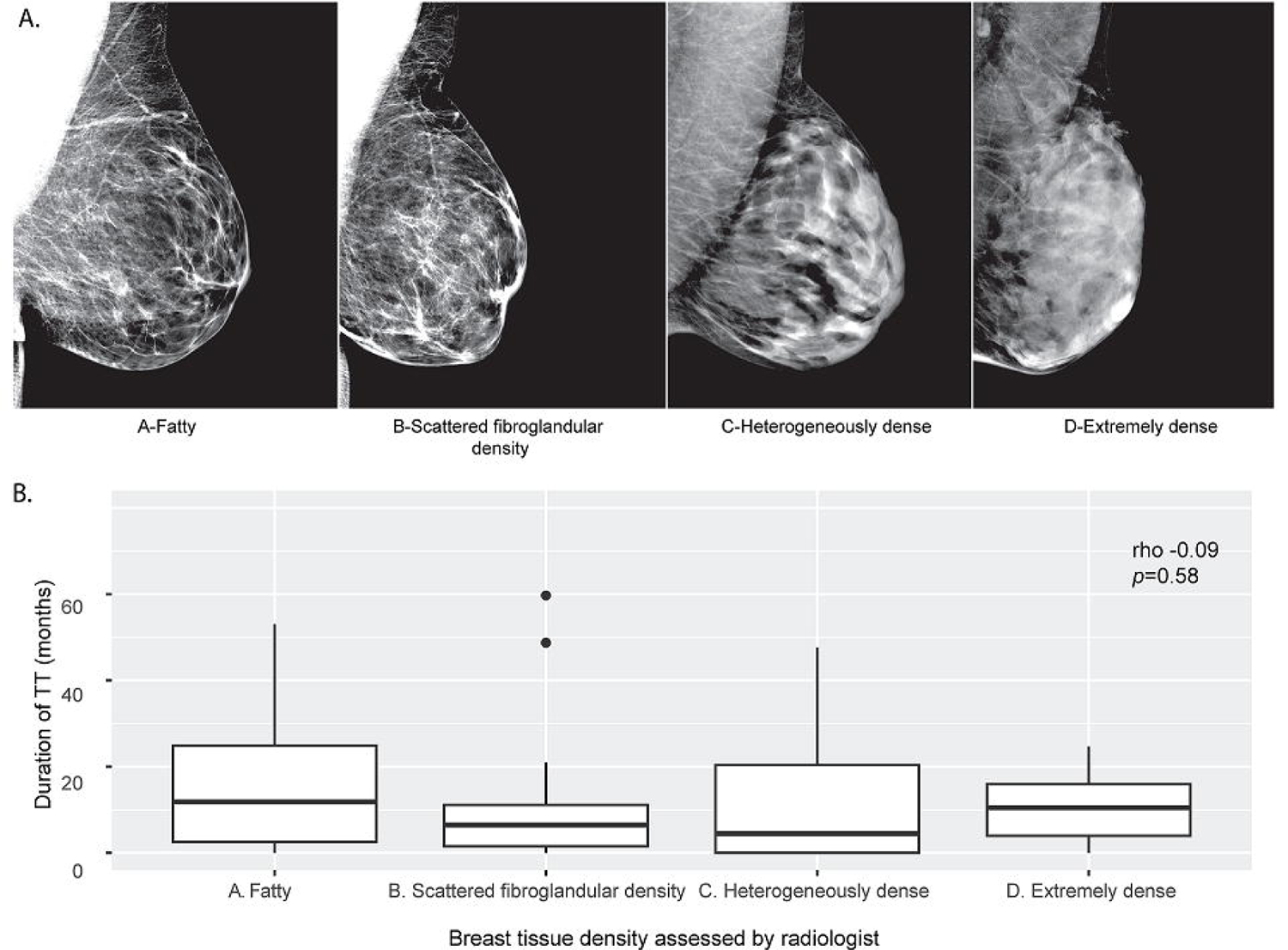
(**A**) Breast tissue density was assessed by a radiologist using four categories with increasing amounts of fibroglandular tissue (epithelium and fibrous stroma). Mammograms were from transmasculine subjects. (**B**) There was no association between the duration of testosterone therapy (TT) and breast tissue density.

### Radiological-pathological correlations

There was no correlation between % epithelium and LIBRA measurements (*p*>0.05; Supplementary 9). However, % fibrous stroma was significantly and positively correlated with both PD (*p*=0.001) and absolute dense area (*p*<0.001), but not with absolute non-dense area (*p*=0.15; Supplementary 9). The amount of fat tissue was significantly inversely correlated with PD (*p*=0.001) and absolute dense area (*p*<0.001), but not with non-dense area (*p*=0.14; Supplementary 9). The results were similar in sensitivity analysis when restricted to individuals who had surgeries within six months after their mammograms (*n*=17; Supplementary 10).

## Discussion

The effect of gender-affirming TT on BC risk is unclear. This study investigated the effect of TT on breast tissue composition in TMIs. In a subset of subjects, we also explored the relationship between TT and mammographic breast density. Leveraging on our deep-learning technology, we demonstrated that longer duration of TT use was associated with decreased amounts of breast epithelium and fibrous stroma. There was no relationship between TT and mammographic breast density. We previously investigated the association of automated breast tissue composition and BC risk in the Nurses’ Health Studies (28). Women in the highest quartile for % epithelium had higher BC risk compared to women in the lowest quartile; there was no relationship between % fibrous stroma and BC risk (28). Taken together, this current study now provides quantitative histological evidence to support prior epidemiological reports that TT may reduce BC risk (3–5).

De Blok *et al* estimated the lifetime BC risk for TMIs to be ≈3.8%, which was lower than the risk in cisgender women (12%) but remained much higher than the risk in cisgender men (<0.1%) (14). BC risk factors for TMIs are not completely established. While it can be assumed that BC risk factors for TMIs are similar to cisgender women (30), their risk can be additionally modified by TT or undergoing chest-contouring surgery. The primary goal of chest-contouring surgery, in contrast to oncologic mastectomies, is the creation of a male-appearing chest rather than the removal of all grossly identifiable breast tissue. In this aspect, chest-contouring surgery resembles reduction mammoplasty. Residual breast tissue after chest-contouring surgery remains hormonally responsive, and BC can still occur (14,34,35). Therefore, even though testosterone is widely reported to have an anti-proliferative effect in the breast (36,37), TMIs who had chest-contouring surgeries retain their inherent BC risk and TT could be modulating that risk. Although the prevalence of atypical lesions and DCIS in our subjects was lower than that observed in cisgender reduction mammoplasties studies (2% vs. 8%) (20,38,39), atypical lesions and DCIS were only found in subjects who received TT compared with non-TT users (23). Our work supports more studies regarding TT in breast pathology, and using preclinical models to understand the extent to which TT affects BC risk in genetically-predisposed individuals (40)

No study has investigated the association between TT and mammographic breast density in the trans masculine population. Therefore, we attempted to gain preliminary insights into the relationship between TT and mammographic breast density. We had limited mammography data because of socioeconomic and biobehavioral challenges face by this population. Excluding young age (<40 years old), mammography is not frequently performed in TMIs due to many factors such as lack of evidence-based screening guidelines, lack of insurance, poor access to medical care, or quite simply, reluctance. It can be emotionally distressing for TMIs to undergo screening for breast and other “female” cancers because of the discordance between their gender identity and their organ inventory, as well as the feminized language around those procedures and being misgendered in the clinics. Nevertheless, the distribution of breast tissue density assessed by the radiologist among the subset of 42 TMIs was similar to the general female population (41). Our radiological-pathological correlations agreed with previous work that mammographic breast density is mostly contributed by fibrous stroma (42,43).

Davis *et al* randomized 250 postmenopausal women to receive placebo, 150 µg/day, or 300 µg/day testosterone transdermal patch, and observed that PD and absolute dense area were not different between paired baseline and week 52 mammograms (44). While the null relationship between TT use and mammographic breast density in our study could be explained by insufficient statistical power, it is possible that the decreases in epithelium and stroma were not large enough to be detected via mammograms. More studies with larger sample sizes and intra-subject mammograms are warranted for definitive insights into the relationship between TT and BC risk. Understanding that relationship will have great clinical impact on BC screening strategies for TMIs and reducing their healthcare disparities.

The strengths of our study include leveraging a large study population with comprehensive pathological review and digital slides to understand trans masculine breast tissue composition as it relates to BC risk. Our study’s limitations include pathologists were not blinded to the gender identity of the population sample which could introduce bias, we were unable to accurately account for TT dosages since dosages are frequently according to how the subject feels (30), limited mammography data, did not collect intra-individual mammograms, and no BMI data at the time of mammography. However, BMI was likely to be similar at mammography and surgery as the time between these two events was relatively short (0.4 to 34.9 months). Lastly, this study was not disaggregated by sex and gender due to the research topic and only consisted of subjects assigned as female at birth.

In conclusion, TT decreases epithelial and fibrous stroma components in breast tissue, supporting epidemiological findings that TMIs receiving TT have lower BC risk compared to cisgender women. More studies are needed to investigate the effect of TT on breast density and BC risk in the trans masculine population.

## Supplementary Data

**Supplementary 1A.** Automated breast tissue composition significantly correlated with pathologists’ assessments. The percentage (%) of epithelium (**A**) significantly inversely correlated with increasing degrees of lobular atrophy (*p*<0.001; Spearman’s rho). Cases where the pathologists classified the stroma as predominantly fibrous or fatty were significantly correlated with higher % of fibrous stroma (**B**; *p*<0.001) or fat (**C**; *p*<0.001), respectively. Each box displays the median, and 25^th^ and 75^th^ percentiles (upper and lower hinges). The lower whisker represents the smallest observation greater than or equal to the lower hinge - 1.5 * inter quartile range (IQR); the upper whisker represents the largest observation less than or equal to upper hinge + 1.5 * IQR.

**Supplementary 1B.** Automated breast tissue composition remained significantly correlated with pathologists’ assessments even when stratified by testosterone therapy (TT) (all *p*<0.001). Spearman’s rho and *p*-values comparing the percentage (%) of each tissue region and pathologists’ assessments are displayed in **A**, **B**, and **C**. Each box displays the median, and 25^th^ and 75^th^ percentiles (upper and lower hinges). The lower whisker represents the smallest observation greater than or equal to the lower hinge - 1.5 * inter quartile range (IQR); the upper whisker represents the largest observation less than or equal to upper hinge + 1.5 * IQR.

**Supplementary 2.** The association of testosterone therapy (per six months duration) and the percentages (%) of each breast tissue region, stratified by body mass index (BMI).

**Supplementary 3.** The association of testosterone therapy (per six months duration) and the percentages (%) of each breast tissue region and among nulliparous subjects.

**Supplementary 4.** Characteristics of 42 transmasculine individuals who had mammography prior to chest contouring surgery.

**Supplementary 5.** Laboratory for Individualized Breast Radiodensity Assessment (LIBRA) breast percent density (PD; **B**), absolute dense area (**C**), and absolute non-dense area (**D**) significantly correlated with the radiologist’s breast tissue density assessment. Each box displays the median, and 25^th^ and 75^th^ percentiles (upper and lower hinges). The lower whisker represents the smallest observation greater than or equal to the lower hinge - 1.5 * inter quartile range (IQR); the upper whisker represents the largest observation less than or equal to upper hinge + 1.5 * IQR.

**Supplementary 6.** The association between testosterone therapy (per six months duration) and Laboratory for Individualized Breast Radiodensity Assessment (LIBRA) measures.

**Supplementary 7.** The association between testosterone therapy (per six months duration) and Laboratory for Individualized Breast Radiodensity Assessment (LIBRA) measures, stratified by body mass index (BMI).

**Supplementary 8.** The association between testosterone therapy (per six months duration) and Laboratory for Individualized Breast Radiodensity Assessment (LIBRA) measures among nulliparous subjects.

**Supplementary 9.** Scatterplot matrix correlating automated breast tissue composition (pink boxes) with Laboratory for Individualized Breast Radiodensity Assessment (LIBRA) measures (grey boxes) among 23 subjects.

**Supplementary 10.** Scatterplot matrix correlating automated breast tissue composition data (pink boxes) with Laboratory for Individualized Breast Radiodensity Assessment (LIBRA) measures (grey boxes) among 17 subjects who had their chest-contouring surgeries within six months of their mammography.

## Data Availability

The source code for our deep learning networks is available at https://github.com/avellal14/BBD_Pipeline. The data that support the findings of this study are available from Dr. Jan Heng.

## Statements

### Transparency declaration

YJH affirms that the manuscript is an honest, accurate, and transparent account of the study being reported. No important aspects of the study have been omitted. The study was carried out as originally planned.

## Author contributions

Conceived and designed the study: YJH GMW. Data analysis: YJH LY. Clinical, pathology, and mammogram data collection: AMT RAB GMB YDG VCB ESM SG PR JP VJF. Computational data acquisition: GV DK VCB YJH. Clinical input: MSI GMW. All authors contributed to the writing and reviewing of the manuscript.

## Acknowledgements

The authors would like to thank the surgical administrative staff for retrieving the surgical case lists; Ashton Black for reviewing the manuscript. The authors assume full responsibility for analyses and interpretation of these data.

