## Supplemental Data for "Effect of testosterone therapy on breast tissue composition and mammographic breast density in trans masculine individuals"

Supplementary 1A

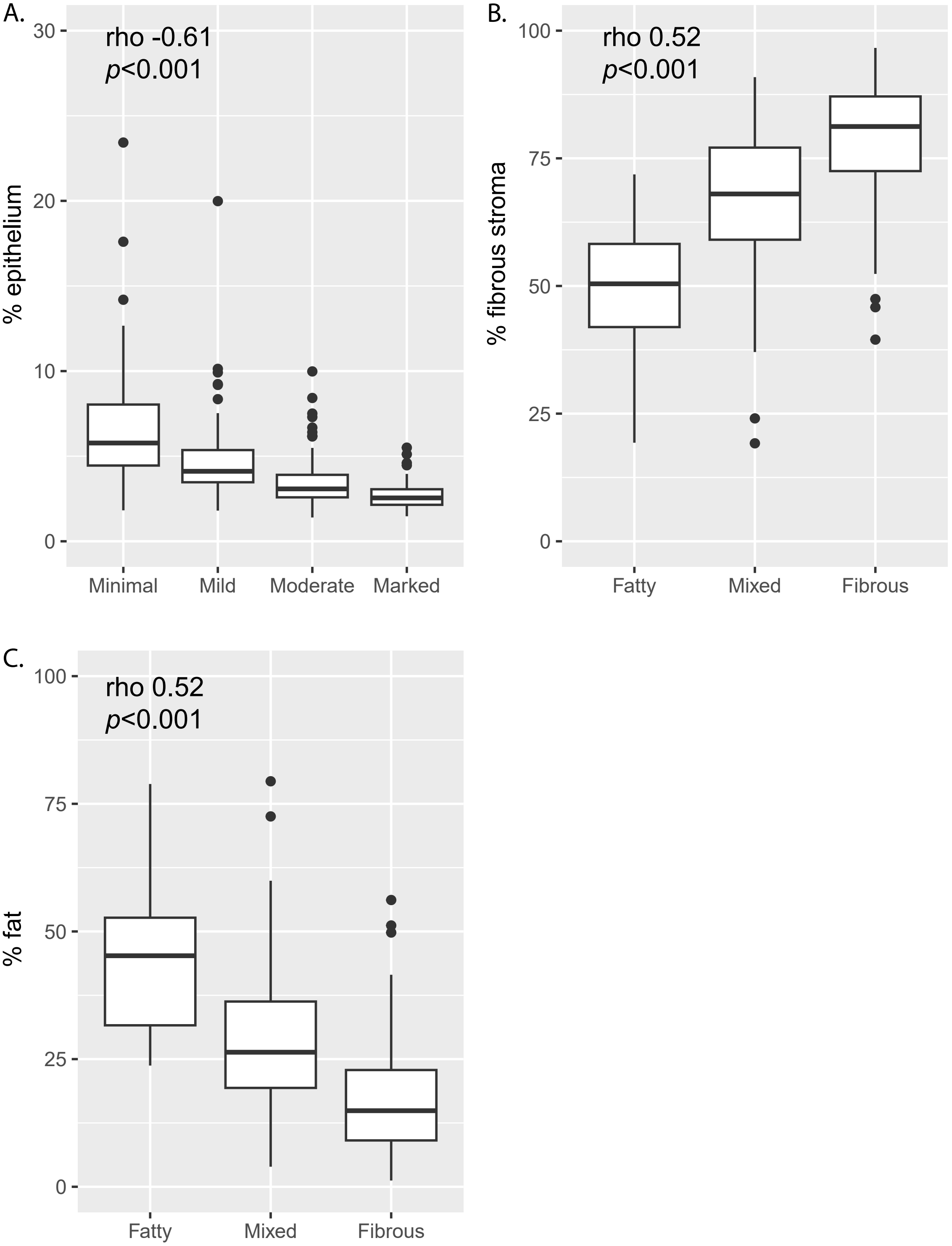

Supplementary 1B

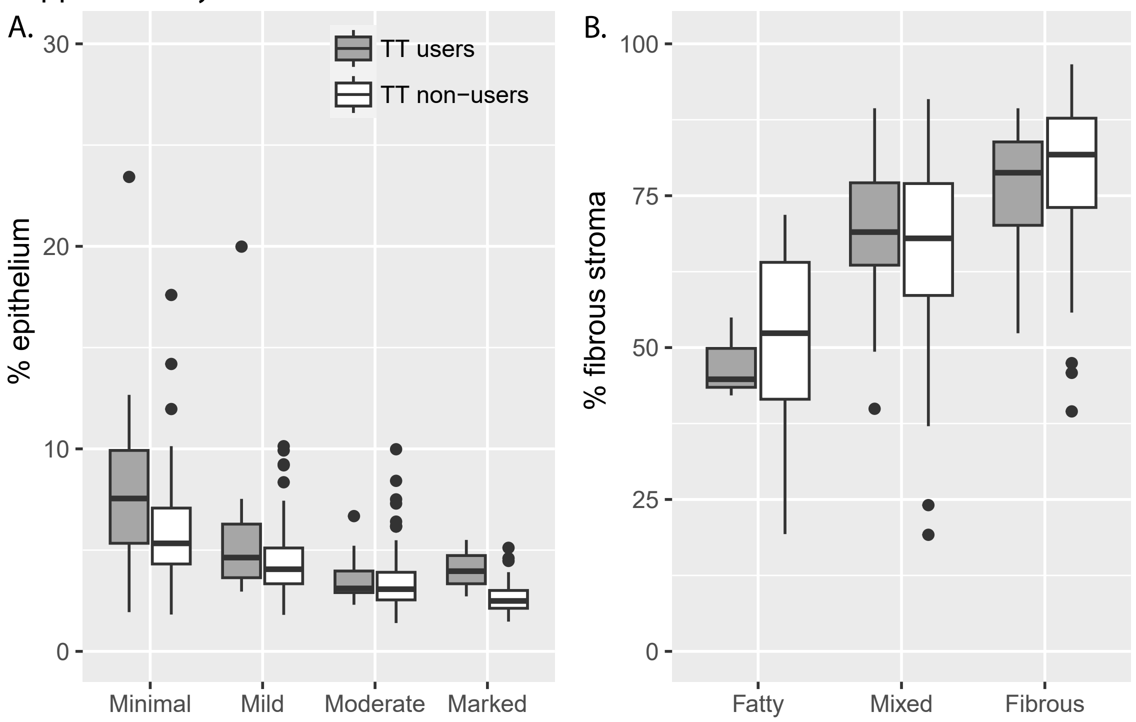


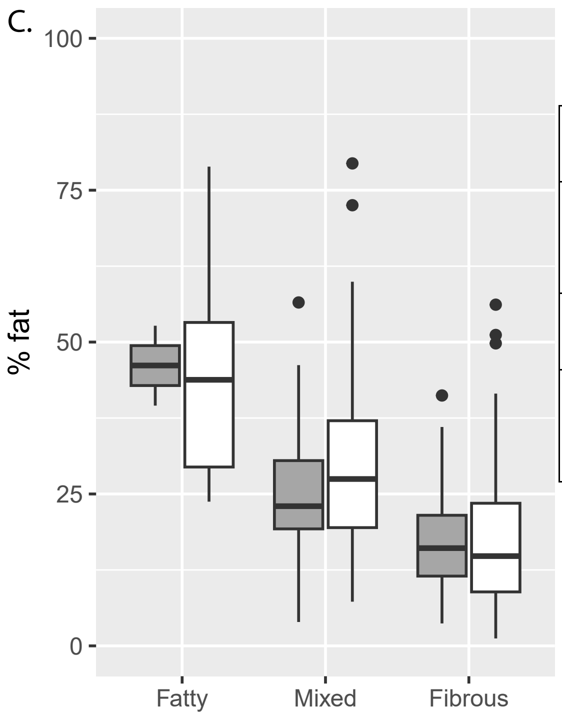


D.

| | TT users | |
|---|---|---|
| | rho | *p* value |
| **% epithelium** | -0.59 | <0.001 |
| **% fibrous stroma** | -0.54 | <0.001 |
| **% fat** | 0.53 | <0.001 |
| | **TT non-users** | |
| | rho | *p* value |
| **% epithelium** | -0.60 | <0.001 |
| **% fibrous stroma** | -0.42 | <0.001 |
| **% fat** | 0.47 | <0.001 |

**Supplementary 2.** The association of testosterone therapy (per six months duration) and the percentages (%) of each breast tissue region, stratified by body mass index (BMI).

| | | *n* | Exp(β) | 95% CI | *p* value |
|---|---|---|---|---|---|
| **BMI <25** | | | | | |
| % epithelium | | | | | |
| | Model 1 | 177 | 0.94 | 0.92-0.97 | <0.001 |
| | Model 2 | 98 | 0.95 | 0.92-0.98 | 0.004 |
| | Model 3 | 91 | 0.96 | 0.93-1.00 | 0.05 |
| % fibrous stroma | | | | | |
| | Model 1 | 177 | 1.00 | 0.99-1.01 | 0.64 |
| | Model 2 | 98 | 1.00 | 0.99-1.01 | 0.60 |
| | Model 3 | 91 | 1.00 | 0.99-1.01 | 0.95 |
| % fat | | | | | |
| | Model 1 | 177 | 1.01 | 0.97-1.05 | 0.54 |
| | Model 2 | 98 | 1.03 | 0.97-1.09 | 0.32 |
| | Model 3 | 91 | 1.02 | 0.96-1.09 | 0.53 |
| **BMI ≥25 and <30** | | | | | |
| % epithelium | | | | | |
| | Model 1 | 134 | 0.98 | 0.96-0.99 | 0.004 |
| | Model 2 | 64 | 0.99 | 0.95-1.02 | 0.44 |
| | Model 3 | 62 | 0.99 | 0.96-1.03 | 0.64 |
| % fibrous stroma | | | | | |
| | Model 1 | 134 | 0.98 | 0.98-0.99 | <0.001 |
| | Model 2 | 64 | 0.99 | 0.98-1.01 | 0.50 |
| | Model 3 | 62 | 1.00 | 0.98-1.01 | 0.69 |
| % fat | | | | | |
| | Model 1 | 134 | 1.02 | 1.00-1.04 | 0.10 |
| | Model 2 | 64 | 1.00 | 0.95-1.04 | 0.85 |
| | Model 3 | 62 | 0.99 | 0.95-1.04 | 0.67 |
| **BMI ≥30** | | | | | |
| % epithelium | | | | | |
| | Model 1 | 98 | 0.97 | 0.95-0.99 | 0.007 |
| | Model 2 | 45 | 0.95 | 0.91-1.01 | 0.08 |
| | Model 3 | 40 | 0.96 | 0.90-1.01 | 0.13 |
| % fibrous stroma | | | | | |
| | Model 1 | 98 | 0.99 | 0.97-1.00 | 0.18 |
| | Model 2 | 45 | 0.97 | 0.94-1.00 | 0.09 |
| | Model 3 | 40 | 0.98 | 0.95-1.00 | 0.10 |
| % fat | | | | | |
| | Model 1 | 98 | 1.01 | 0.98-1.04 | 0.36 |
| | Model 2 | 45 | 1.04 | 0.97-1.13 | 0.26 |
| | Model 3 | 40 | 1.01 | 0.94-1.09 | 0.68 |

Model 1 adjusted for age and year of surgery. Model 2 adjusted for age and year of surgery, race/ethnicity, chest binding, and oophorectomy status. Model 3 adjusted for age and year of surgery, race/ethnicity, chest binding, oophorectomy status, and estimated weekly testosterone dose. Confidence interval, CI.

**Supplementary 3.** The association of testosterone therapy (per six months duration) and the percentages (%) of each breast tissue region among nulliparous subjects.

| | | ***n*** | **Exp(β)** | **95% CI** | ***p* value** |
|---|---|---|---|---|---|
| % epithelium | | | | | |
| | Model 1 | 171 | 0.97 | 0.95-0.98 | 0.001 |
| | Model 2 | 87 | 0.96 | 0.93-0.99 | 0.02 |
| | Model 3 | 82 | 0.96 | 0.92-0.99 | 0.02 |
| % fibrous stroma | | | | | |
| | Model 1 | 171 | 1.00 | 0.99-1.01 | 0.75 |
| | Model 2 | 87 | 0.99 | 0.98-1.01 | 0.27 |
| | Model 3 | 82 | 0.99 | 0.98-1.01 | 0.29 |
| % fat | | | | | |
| | Model 1 | 171 | 1.01 | 0.98-1.04 | 0.49 |
| | Model 2 | 87 | 1.02 | 0.98-1.07 | 0.32 |
| | Model 3 | 82 | 1.01 | 0.97-1.06 | 0.49 |

Model 1 adjusted for age and year of surgery. Model 2 adjusted for age and year of surgery, race/ethnicity, chest binding, and oophorectomy status. Model 3 adjusted for age and year of surgery, race/ethnicity, chest binding, oophorectomy status, and estimated weekly testosterone dose. Confidence interval, CI.

**Supplementary 4.** Characteristics of 42 transmasculine individuals who had mammography prior to chest contouring surgery.

| | All Individuals | Individuals with DICOM files | Individuals without DICOM files | *p* value |
|---|---|---|---|---|
| **N (%)** | 42 | 25 | 17 | |
| **Age at mammogram, median [IQR]** | 43.3 [37.8, 48.5] | 38.8 [30.6, 45.5] | 46.1 [39.5, 49.3] | 0.09[a] |
| **Race/ethnicity, *n* (%)** | | | | 0.35[b] |
| White | 33 (78.6) | 18 (72.0) | 15 (88.2) | |
| Black or African American | 6 (14.3) | 5 (20.0) | 1 (5.9) | |
| Asian | 1 (2.4) | 0 (0.0) | 1 (5.9) | |
| Multiracial | 1 (2.4) | 1 (4.0) | 0 (0.0) | |
| Native American/Pacific Islander | 1 (2.4) | 1 (4.0) | 0 (0.0) | |
| **Family history of breast cancer, *n* (%)** | | | | 0.33[b] |
| Yes | 13 (31.0) | 9 (36.0) | 4 (23.5) | |
| No | 27 (64.3) | 14 (56.0) | 13 (76.5) | |
| Not reported | 2 (4.8) | 2 (8.0) | 0 (0.0) | |
| **Oophorectomy prior to mammogram, *n* (%)** | | | | 1.00[b] |
| Yes | 5 (11.9) | 3 (12.0) | 2 (11.8) | |
| No | 37 (88.1) | 22 (88.0) | 15 (88.2) | |
| **BMI at surgery, median [IQR]** | 28.5 [24.6, 30.0] | 28.7 [25.7, 30.1] | 26.5 [24.1, 29.8] | 0.51[a] |
| **Duration of testosterone therapy at mammogram, *n* (%)** | | | | 0.45[b] |
| Never | 9 (21.4) | 3 (12.0) | 6 (35.3) | |
| <1 year | 8 (19.0) | 5 (20.0) | 3 (17.6) | |
| >=1 to <2 years | 15 (35.7) | 10 (40.0) | 5 (29.4) | |
| >=2 to <5 years | 9 (21.4) | 6 (24.0) | 3 (17.6) | |
| >=5 years | 1 (2.4) | 1 (4.0) | 0 (0.0) | |
| **Chest binding at mammogram, *n* (%)** | | | | 1.00[b] |
| Yes | 19 (45.2) | 16 (94.1) | 3 (100.0) | |
| No | 1 (2.4) | 1 (5.9) | 0 (0.0) | |
| Not reported | 22 (52.4) | 3 (12.0) | 3 (17.6) | |
| **Breast tissue density assessed by the radiologist** | | | | 0.48[b] |
| A-Fatty | 6 (14.3) | 3 (12.0) | 3 (17.6) | |
| B-Scattered fibroglandular | 13 (31.0) | 7 (28.0) | 6 (35.3) | |
| C-Heterogeneously dense | 18 (42.9) | 13 (52.0) | 5 (29.4) | |
| D-Dense | 5 (11.9) | 2 (8.0) | 3 (17.6) | |

*p*-values were obtained using the [a]Mann-Whitney or [b]Fisher's exact test by comparing those that had DICOM files versus those that did not. Data in "not reported" categories were excluded from statistical analysis. Body mass index, BMI; Inter-quartile range, IQR. Percentages may not add up to 100% due to rounding.

Supplementary 5

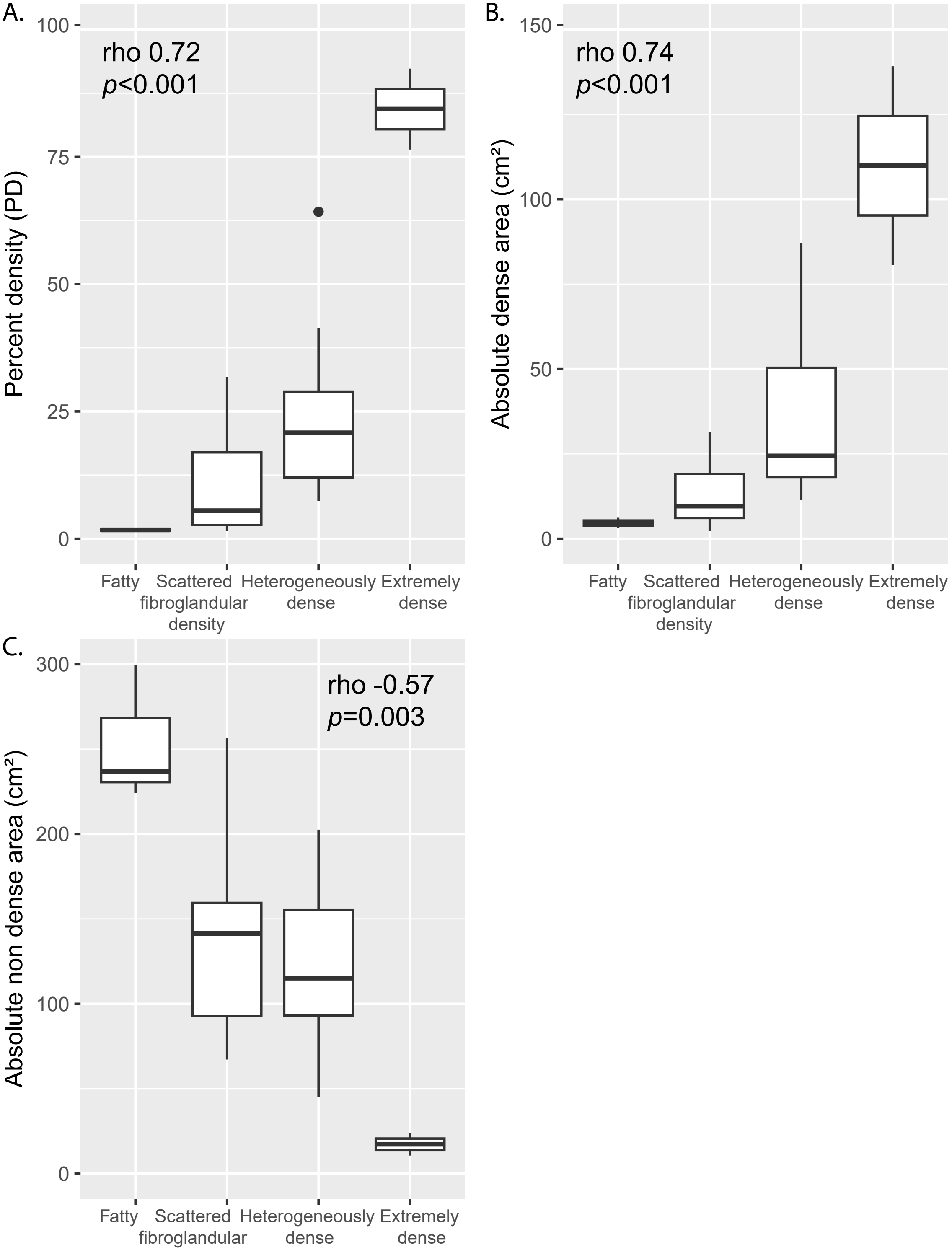

**Supplementary 6.** The association between testosterone therapy (per six months duration) and LIBRA measures.

| | *n* | Exp(β) | 95% CI | *p* value |
|---|---|---|---|---|
| **Percent density** | | | | |
| Crude | 25 | 0.89 | 0.76-1.06 | 0.18 |
| Adjusted model | 25 | 0.97 | 0.83-1.14 | 0.71 |
| **Absolute dense area** | | | | |
| Crude | 25 | 0.90 | 0.77-1.04 | 0.14 |
| Adjusted model | 25 | 0.94 | 0.81-1.09 | 0.41 |
| **Absolute non-dense area** | | | | |
| Crude | 25 | 1.05 | 0.95-1.17 | 0.31 |
| Adjusted model | 25 | 1.00 | 0.92-1.08 | 0.92 |

The adjusted model included age at mammogram and BMI at chest-contouring surgery.

**Supplementary 7.** The association between testosterone therapy (per six months duration) and LIBRA measures, stratified by body mass index (BMI).

| | *n* | Exp(β) | 95% CI | *p* value |
|---|---|---|---|---|
| **BMI ≤25** | | | | |
| **Percent density** | | | | |
| Crude | 6 | 1.06 | 0.68-1.65 | 0.74 |
| Adjusted model | 6 | 1.09 | 0.67-1.78 | 0.61 |
| **Absolute dense area** | | | | |
| Crude | 6 | 0.96 | 0.63-1.47 | 0.80 |
| Adjusted model | 6 | 0.98 | 0.60-1.60 | 0.93 |
| **Absolute non-dense area** | | | | |
| Crude | 6 | 0.92 | 0.67-1.27 | 0.52 |
| Adjusted model | 6 | 0.90 | 0.64-1.26 | 0.40 |
| **BMI>25** | | | | |
| **Percent density** | | | | |
| Crude | 19 | 0.89 | 0.74-1.08 | 0.23 |
| Adjusted model | 19 | 0.88 | 0.72-1.08 | 0.21 |
| **Absolute dense area** | | | | |
| Crude | 19 | 0.90 | 0.75-1.08 | 0.23 |
| Adjusted model | 19 | 0.88 | 0.73-1.07 | 0.19 |
| **Absolute non-dense area** | | | | |
| Crude | 19 | 1.06 | 0.94-1.18 | 0.32 |
| Adjusted model | 19 | 1.08 | 0.96-1.21 | 0.18 |

The adjusted model included age at mammogram only.

**Supplementary 8.** The association between testosterone therapy (per six months duration) and LIBRA measures among nulliparous subjects.

| | *n* | Exp(β) | 95% CI | *p* value |
|---|---|---|---|---|
| **Percent density** | | | | |
| Crude | 11 | 0.82 | 0.64-1.05 | 0.10 |
| Adjusted model | 11 | 0.88 | 0.67-1.16 | 0.31 |
| **Absolute dense area** | | | | |
| Crude | 11 | 0.82 | 0.66-1.02 | 0.07 |
| Adjusted model | 11 | 0.86 | 0.67-1.11 | 0.21 |
| **Absolute non-dense area** | | | | |
| Crude | 11 | 1.08 | 0.90-1.29 | 0.37 |
| Adjusted model | 11 | 1.06 | 0.92-1.22 | 0.35 |

The adjusted model included age at mammogram and BMI at chest-contouring surgery.

Supplementary 9

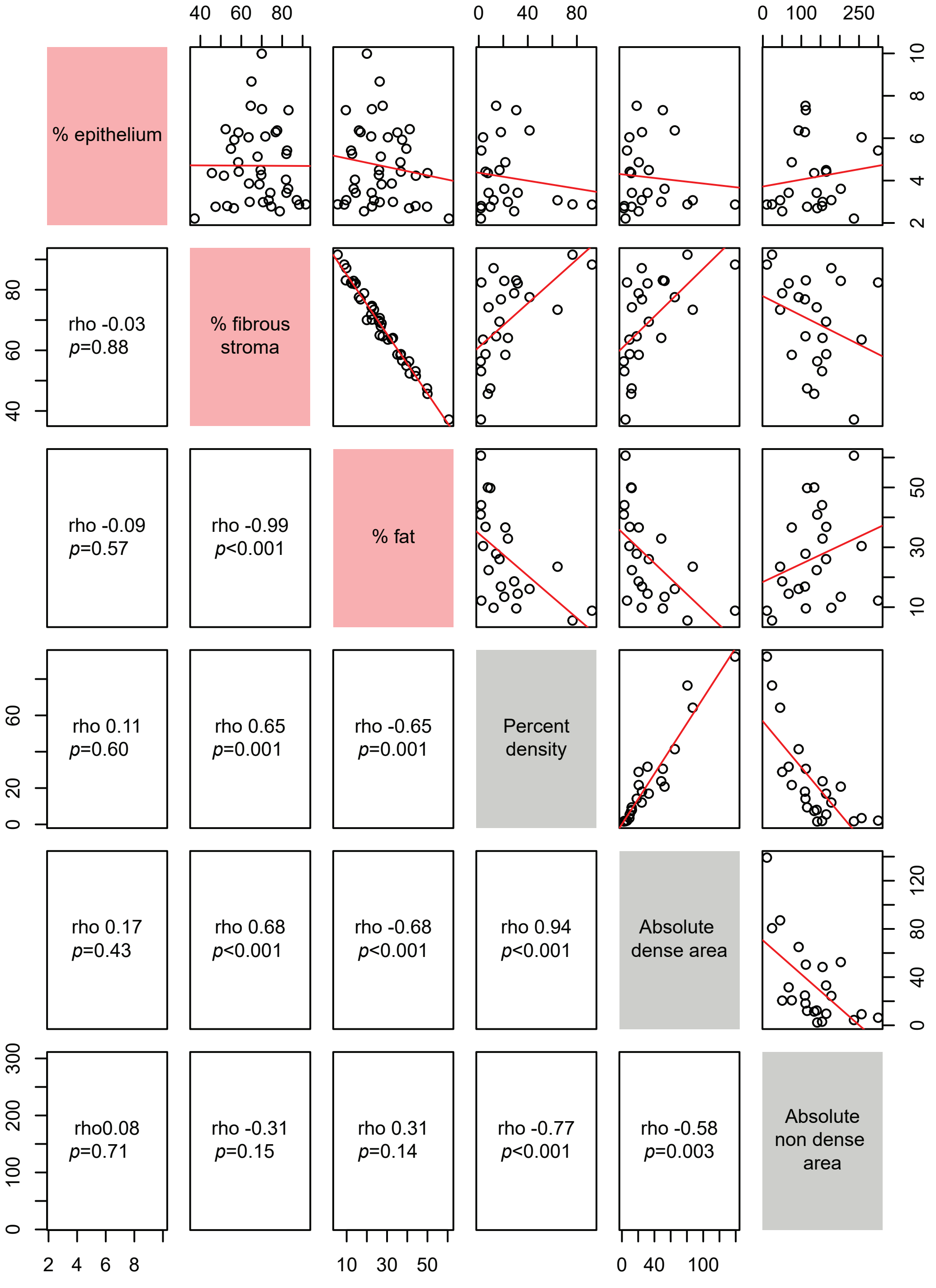

### Supplementary 10

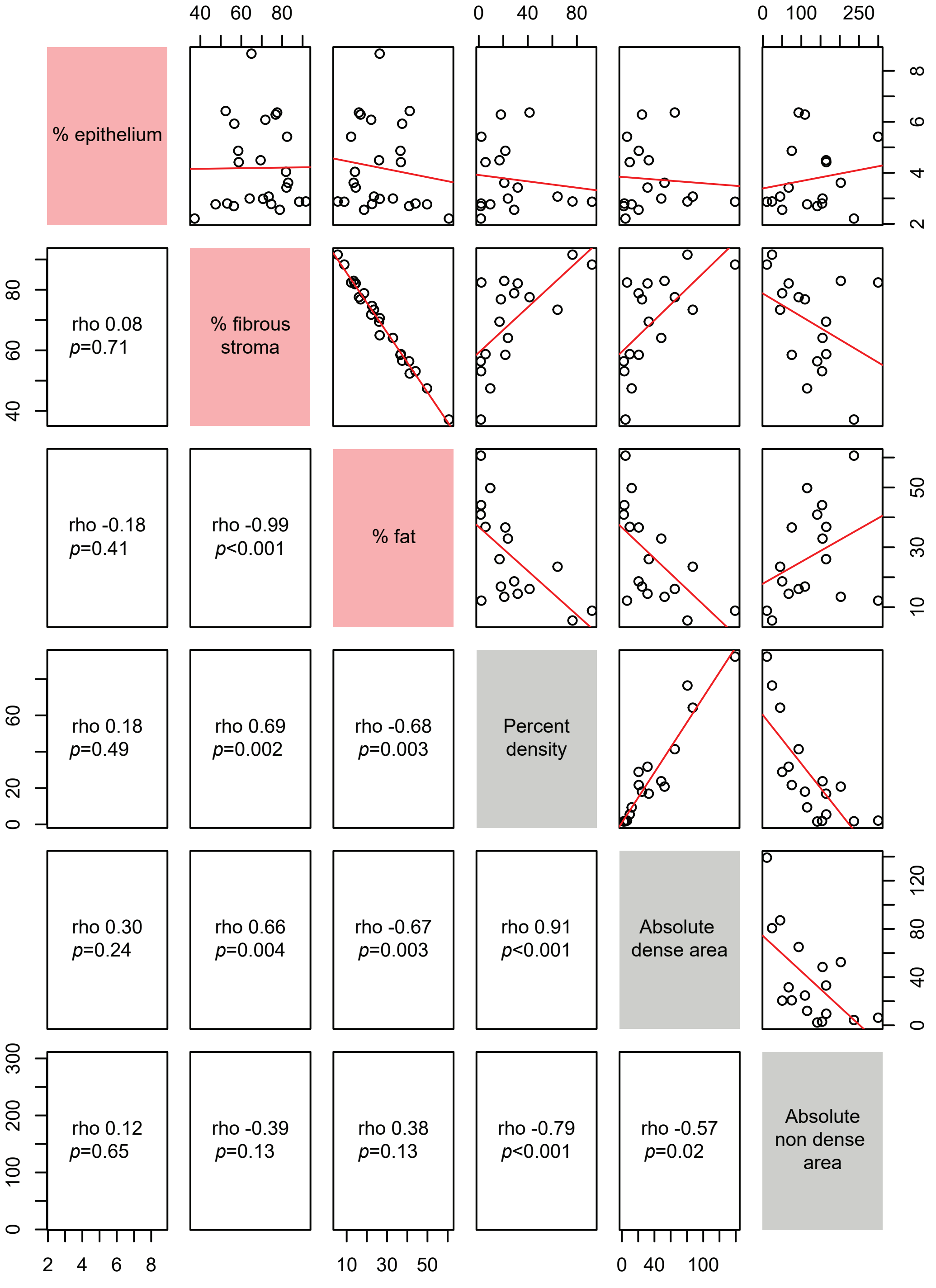
